# AI Clinical Decision Support System (CDSS) for Teleconsultations in eSanjeevani, India Telemedicine Service: A Prospective Implementation Study

**DOI:** 10.1101/2025.11.22.25340800

**Authors:** Prasan Kumar Panda, Tonmoy Dutta, Ashish Chander, Satish Kalikar, Shreya Thakur, Sanjay Sood, Sumandeep Singh, Gagandeep Singh, Davinder Bisht, Kanhaiya Lal, Amit Kumar Tyagi, Mudita Khattri, Meenu Singh

## Abstract

**Background:** Most Western clinical decision support systems (CDSS) fail to contextualise to India’s healthcare realities, limiting their adoption by healthcare workers (HCWs) and patients. With the rapid evolution of India’s telemedicine ecosystem, there is an urgent need for an indigenous, validated, and scalable AI-based CDSS integrated within national platforms to improve diagnostic accuracy and care delivery.

**Methods:** This study, conducted between 2022–2024 by an AI Centre of Excellence of the Government of India, focused on developing, validating, and implementing a knowledge-based CDSS symptom entry Physician Assistance Form (PAF) within eSanjeevani—India’s national teleconsultation platform. The study was conducted in three phases, each of which was interdependent, overlapping, and dynamic in nature.

**Phase 1:** The AI system development utilised retrospective eSanjeevani 1.0 data (64 million consultations), extracting 0.22 million SNOMED CT–aligned records to identify 29,000 unique symptoms, which were refined to 115 for model training in the initial plan during 2022. The upgraded eSanjeevani 2.0 (2023) incorporated AI-based differential diagnosis, which was later expanded to include 300 symptoms (2025) with branching logic that integrated patient age, gender, language, and multidimensional symptom attributes.

**Phase 2:** Expert clinicians validated the symptom repository, logic flow, and AI-generated diagnoses.

**Phase 3:** The validated CDSS was implemented in eSanjeevani 2.0, providing real-time differential diagnosis and departmental recommendations during assisted and non-assisted teleconsultations.

**Findings:** Integration of AI-CDSS improved structured data capture, enhanced diagnostic precision, and streamlined patient triage within teleconsultations.

**Interpretation:** India’s AI-CDSS initiative represents the first government-supported, large-scale CDSS integration in a developing country, offering a replicable, ethical, and contextually grounded model for other LMICs to advance equitable and quality telehealth services.

## Introduction

The World Health Organization (WHO) advocates a minimum doctor: population ratio of 1:1000. Although this prescribed ratio is present in most of the Western world, 44% of the WHO member states report less than 1 physician per 1,000 patients [1]. In 2022, India had 0.8 medical doctors per 1,000 population, though this ratio is rapidly changing [2]. Additionally, there is a shortage of trained manpower in the healthcare system, with nurses at 1.7 per 1,000 people and pharmacists at 0.8 per 1,000 people. In rural India, where the number of primary health care centers (PHCs) is limited, 8% of the centers do not have doctors or medical staff, 39% do not have lab technicians and 18% PHCs do not even have a pharmacist. More importantly, there is a shortfall of 24% sub-centers, 29% PHCs and 38% CHCs across the country as per the Rural Health Statistics (RHS) 2020 [3]. The situation remains worrisome in rural areas, where almost 66 percent of India’s population resides. Being a developing country, India can’t have immediate solutions to these crises. However, the COVID-19 pandemic and the growing number of digital users have led to rapid solutions for various aspects, including teleconsultation. Furthermore, to address these acute shortages and to assist the existing doctors/staffs/patients in the healthcare system, artificial intelligence (AI) is rapidly evolving.

The integration of AI into telemedicine has shown potential to enhance clinical decision-making by supporting physicians in real-time consultations. eSanjeevani, India’s national telemedicine platform of MoHFW, Government of India, has evolved into the world’s largest documented telemedicine implementation in primary healthcare [4]. The platform facilitates between 0.4 and 0.45 million consultations per day. It has witnessed a rapid surge in consultations, necessitating AI-driven solutions to improve diagnostic accuracy and workflow efficiency. Clinical decision support systems (CDSS) powered by AI can assist in symptom evaluation, differential diagnosis generation, and department recommendations, reducing variability in clinical judgment and streamlining patient management.

CDSS are computer programs that assist healthcare professionals in augmenting clinical decision-making processes. These systems are designed to provide healthcare professionals with actionable insights, evidence-based recommendations, and patient-specific information at the point of care, thereby enhancing diagnostic accuracy, treatment efficacy, and patient outcomes. It operates on a foundation of medical knowledge, algorithms, and patient data, aiming to bridge the gap between vast medical information and timely, informed clinical decisions [5]. However, expert-driven validation is essential to ensure accuracy and clinical relevance before implementation in real-world settings [6,7]. Although CDSS began in the 1970s, its real utility in developed countries did not emerge until the 2000s. It has been proven again and again, with its success stories, pitfalls, and potential harms [8].

The primary issue with India’s use case in relation to the available Western CDSS model is its lack of contextualization for local settings. And the local population, including healthcare workers (HCWs), have shown the greatest inertia to kick start the use. However, due to the growing dynamic need of India’s healthcare landscape, there is a need for ongoing development and validation of its own AI-based CDSS in local settings. Both patients and healthcare workers need this CDSS assistance for rapid and high-quality care delivery. To start with, OPD teleconsultation can serve as a platform for utilising CDSS to capture accurate data, achieve precise diagnostics, and facilitate scientific patient triage.

Hence, this study focuses on the development, validation, and implementation of a knowledge-based CDSS symptom entry physician assistance form (PAF) for eSanjeevani-assisted and non-assisted teleconsultations, predicting the correct diagnosis, recommending the appropriate department to patients if required, and thus improving the model.

## Methodology

This was a prospective AI-CDSS implementation study conducted from 2022 to 2024 by one of the AI centres of excellence (COE) of the Government of India, namely AIIMS Rishikesh, in collaboration with its AI technical partner, Wadhwani AI, and data provider and deployment partner, CDAC Mohali. eSanjeevani teleconsultation was initiated in 2020, utilising the initial version, eSanjeevani 1.0, to record patient complaints/symptoms in a free-text format. The data, thus generated, were unstructured and unclassified, resulting in inconsistencies in patient complaint documentation, which made interpretation difficult during remote teleconsultations. To assist these consultations in the right direction, CDSS model was planned in three phases, and each phase was concurrent with time, overlapping, and progressive in nature:

**Phase 1: AI system development (Fig. 1)**

**Figure 1:**
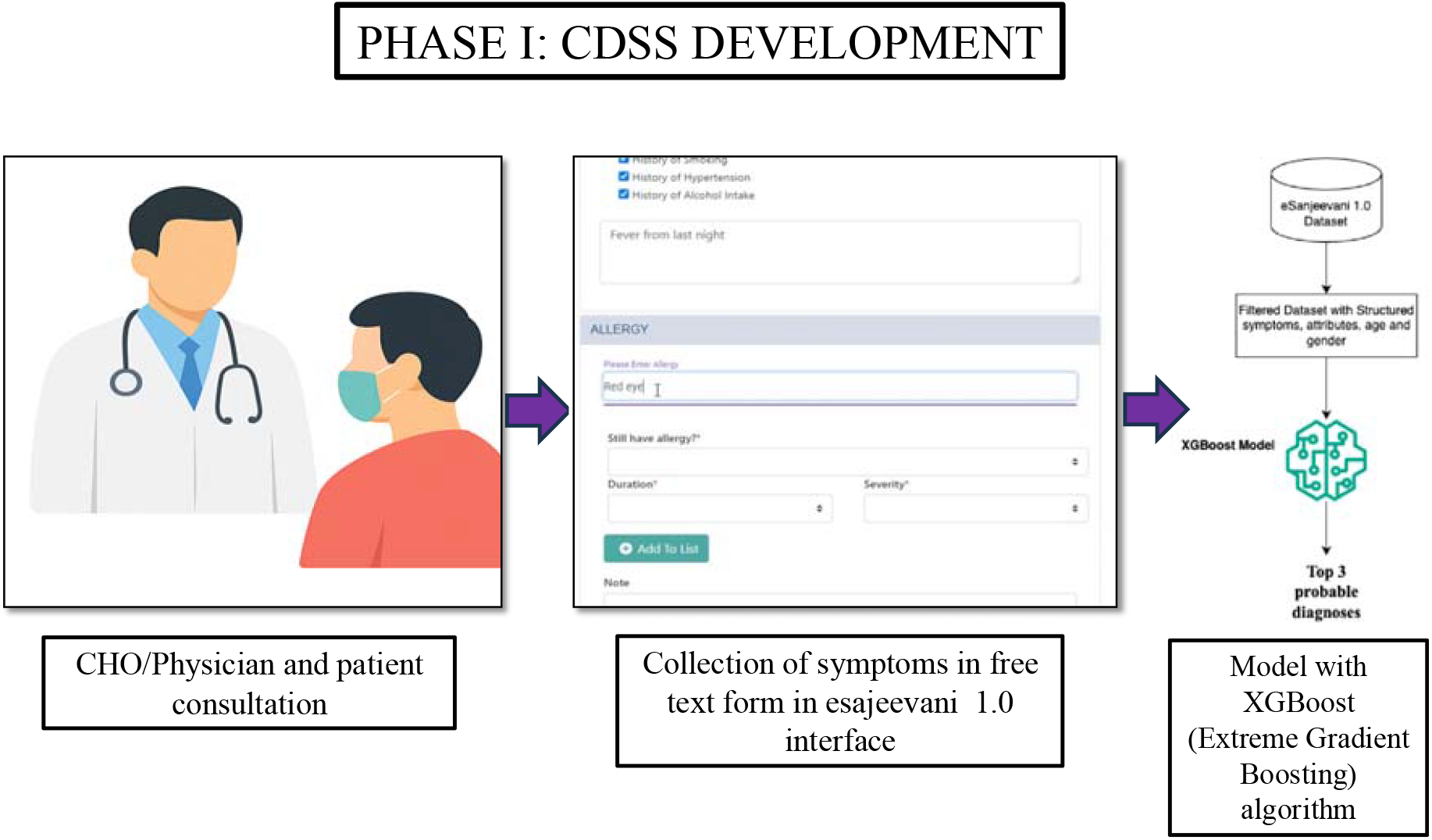
Phase I Development: Physician/ Community health officer (CHO) consultation with collection of data in free text form in eSanjeevani 1.0 to create a physician-assisted form (PAF).

### Data collection & feature engineering

Teleconsultation records from eSanjeevani 1.0 were extracted in 2022 and subsequently standardized as per SNOMED CT medical coding. From these consultations, symptoms were identified based on unique symptom counts/records, following which the data were further filtered for analysis.

To define the attributes of each symptom, sequential questions were designed to gather detailed information based on various guidelines published by national authorities for patient care, as well as WebMD, medical textbooks, and the CDC. Each question sequencing was based on onset, location, duration, characteristics, aggravating factors, relieving factors, and timing (OLDCART) [9]. Ultimately, a symptom repository was created.

### Model training

AI algorithms were trained using labelled consultation data from eSanjeevani 1.0, where each record represented a completed doctor–patient consultation. The dataset included structured symptom information along with relevant demographic and clinical attributes. XGBoost (Extreme Gradient Boosting) algorithm was selected for model development.

Each record in the training dataset contained:

- Input features: structured symptoms with associated attributes such as duration, severity, and onset, along with age and gender.
- Target variable: the doctor-confirmed diagnosis recorded in eSanjeevani 1.0.

The learning task was formulated as a multi-class classification problem, where the model predicts the most likely disease from a predefined set of possible outcomes.

After successful training and validation, the XGBoost model was integrated into the CDSS backend service. When a patient’s symptoms are recorded through the physician assistance form (PAF) in eSanjeevani, the captured data is pre-processed and passed to the deployed model for real-time inference.

The model then generates the Top 3 probable diagnoses along with their corresponding confidence scores. These AI-generated outputs are displayed on the remote doctor’s interface within eSanjeevani, enabling consistent, evidence-based decision-making and ensuring standardized care delivery across Health and Wellness Centres.

### Prototype development

**A** community health officer (CHO)/physician-assisted form (PAF) with a user-friendly interface for telemedicine consultants was designed to capture patient symptoms and attributes in a structured, SNOMED-aligned format prior to teleconsultation. The PAF predicted DDs and Dept. recommendation.

**Phase 2: Expert knowledge-based validation (Fig. 2)**

**Figure 2:**
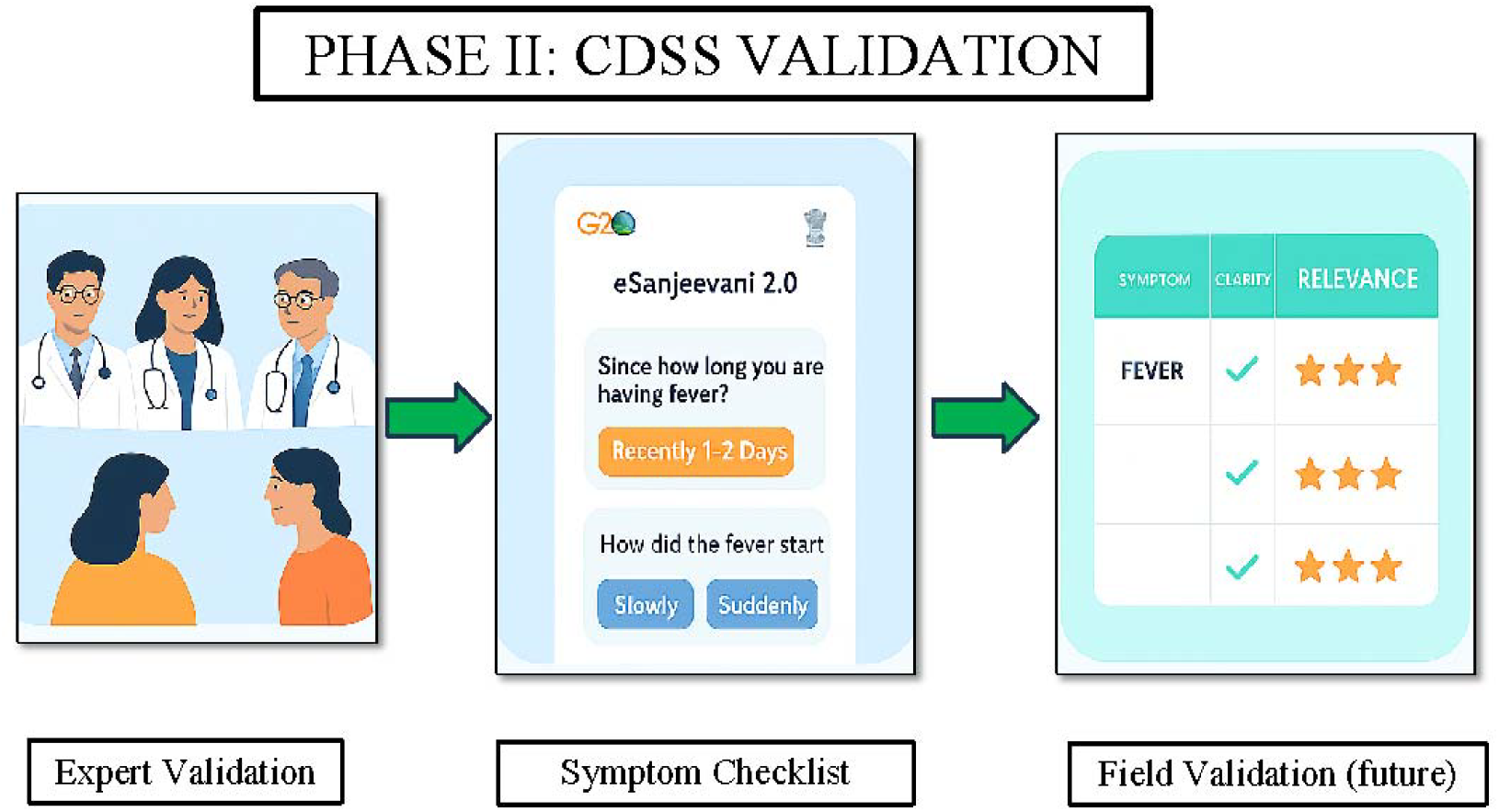
Phase II Validation: Three independent experts validating the physician-assisted form (PAF) along with its branching questionnaire, with subsequent refinement based on dynamic feedback. Once it’s used in teleconsultation, it undergoes field validation through this feedback mechanism.

### Study experts

The CDSS PAF was evaluated by three independent medical experts in clinical decision-making in 2023. All experts were consultant physicians from the various governmental institutes of national importance.

### Validation process

Experts assessed symptom repository, branching logic, and AI-generated differential diagnoses and department recommendations. Each symptom and its corresponding questionnaire were validated based on clarity, relevance, patient and user satisfaction, and logical flow. Multiple rounds of expert feedback and refinement were conducted over a three-month period.

### Multilingual adaptation

The validated symptom repository was translated into 12 regional Indian languages for region-specific implementation in eSanjeevani 2.0. These included Hindi, Tamil, Telugu, Malayalam, Kannada, Marathi, Odia (Oriya), Punjabi, Gujarati, Assamese, Bengali, and Urdu.

**Phase 3: Implementation (Fig. 3)**

**Figure 3:**
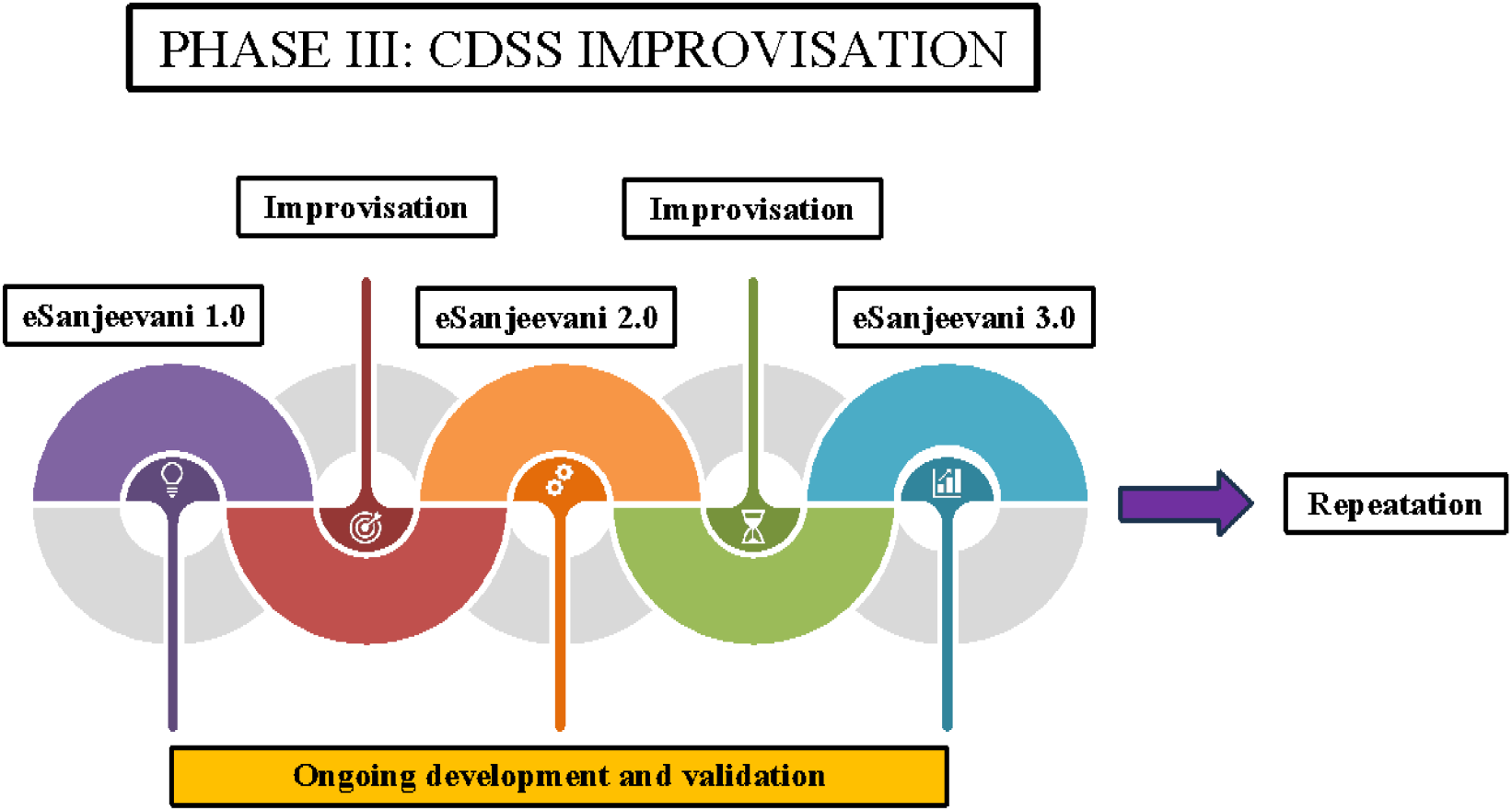
Phase III Improvisation: The ongoing cycle of validation, implementation, and improvisation, highlighting the continuous development of newer model versions.

### Deployment

In March 2023, the validated CDSS-PAF, comprising the clinical symptom repository and AI-generated differential-diagnosis support, was integrated into the eSanjeevani 2.0 platform.

### Evaluation metrics

Feedback from the CHO and Physician of the spoke and hub, respectively, was collected to evaluate awareness, usability, clinical relevance, and satisfaction with the CDSS. Additionally, the effectiveness of the AI-generated differential diagnosis and department recommendations was systematically evaluated.

### Health-system impact

Structured symptom documentation was generated using PAF as per SNOMED CT terminology.

### Continuous improvement

The PAF and symptom workflows have undergone iterative refinement over time, incorporating additional data and continuous user feedback.

#### Ethical and statistics considerations

The study was conducted in accordance with ethical guidelines and obtained institutional ethics committee approval (431/IEC//lM/NF/2022). Data privacy and confidentiality measures were implemented. Considering the nature of the data, only descriptive statistics were used, encompassing both numerical and qualitative data.

## Results

This national, structured, phased, CDSS (PAF & AI generated differential diagnosis support to physicians and CHO) in eSanjeevani teleconsultation was successfully developed, validated and deployed (Figure 4). eSanjeevani daily teleconsultations have been increasing day by day and reached >250million consultations at the end of year 2024 (Figure 5).

**Figure 4:**
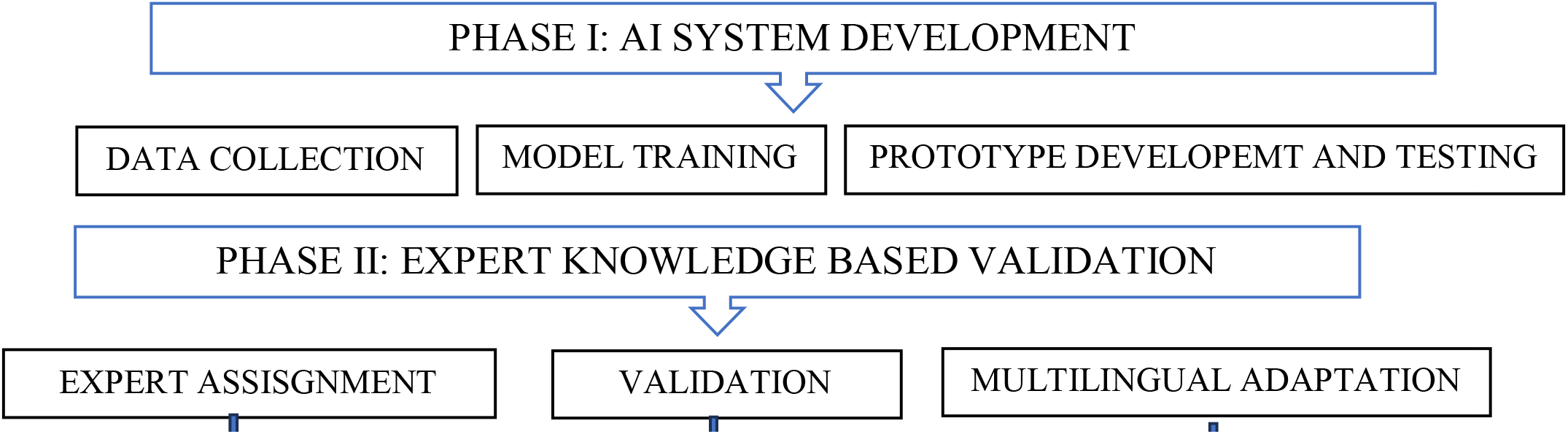
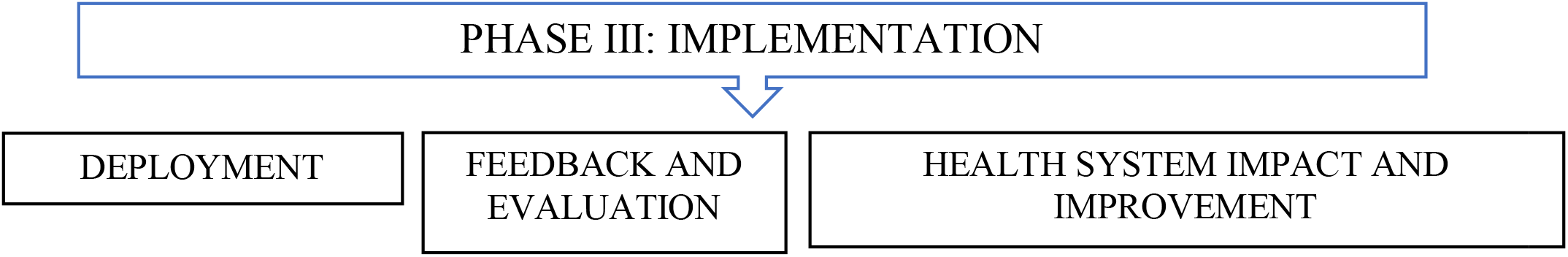
Study flow diagram.

**Figure 5:**
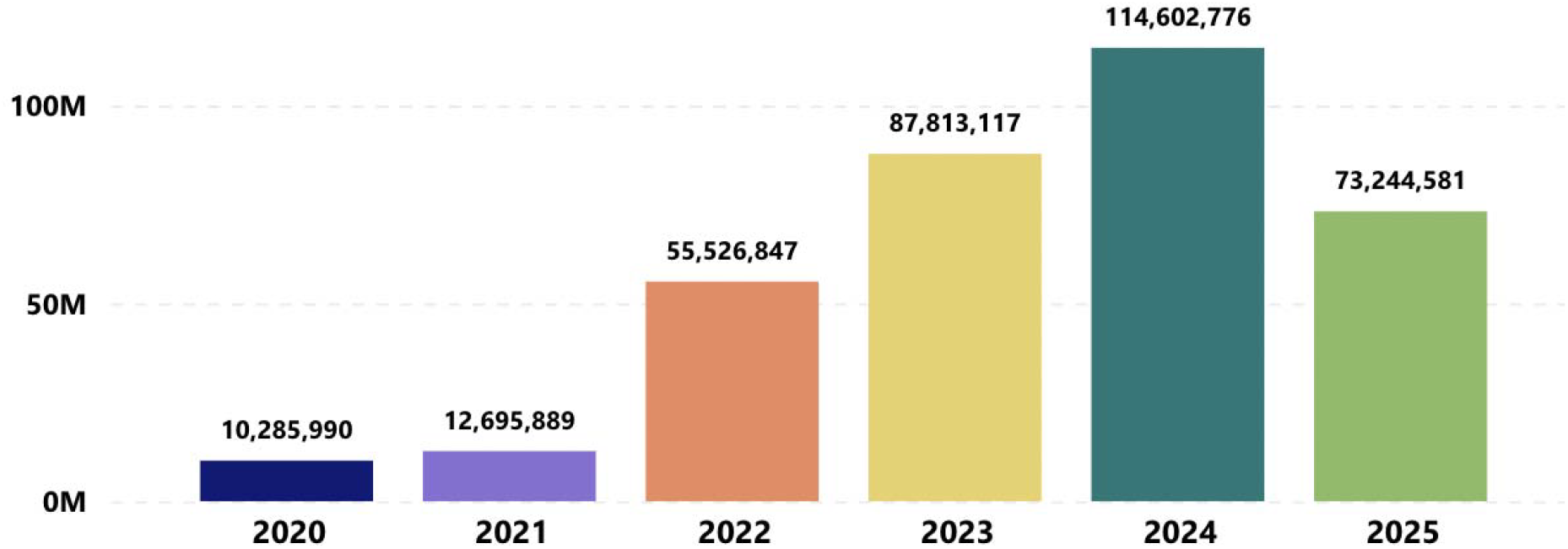
Yearwise teleconsultations in eSanjeevani till today.

### AI system development

Retrospective data from eSanjeevani 1.0 consultation record of 64 million data, only 0.22 million consultation data that aligned with SNOMED CT medical coding were extracted. Among these consultation data based on unique symptom count, 29,000 symptoms were extracted, which were further filtered into 115 medically defined symptoms and created PAF and AI-based differential diagnosis (30) was predicted using symptoms only. Later on, the model integrated 300 symptoms, considering the attributes of symptom details, patient age, and gender, to predict differential diagnosis. These symptoms included 14 disorders/chronic conditions, 285 clinical symptoms, and one ‘other’ category of non-available symptom (Supplementary table). Thirty-one DDs were created with the 17 OPD recommendations. The average no of attributes for a symptom/disease was six, with a minimum of 2 and a maximum of 20.

### Expert knowledge-based validation

The 300 symptoms repository or PAFs was validated using expert opinion, where they assessed the symptom repository, logic flow of the branching questionnaire, and AI-generated differential diagnosis through shared Excel. Independently, they validated each symptom and its attributes and later on agreed with consensus (Kappa, 0.92).

### Implementation

The validated CDSS (115 symptom workflows & AI generated 30 differential diagnosis support to doctors) was deployed in eSanjeevani 2.0 in March 2023, enabling CHO and physicians to record symptoms and receive AI support in real-time. Later in January 2025, an updated version 2.0 of CDSS (300 symptom workflows) was integrated with eSanjeevani, considering the attributes of symptom details, patient age, and gender in predicting differential diagnosis among 31 diseases and 17 OPD recommendations. Also, eSanjeevani 2.0 had been integrated into teleconsultation services in above 12 Indian languages. Recently, two additional languages, Manipuri and Konkani, were added to the symptom repository CDSS.

### Optimisation

To achieve continuous improvement in this CDSS, real-life feedback was incorporated through the eSanjeevani portal from both the CHO/physician and patient sides, allowing for ongoing changes over time. In June 2025, a survey was conducted among CHO and physicians who were using the eSanjeevani 2.0 teleconsultation platform across India to understand awareness, usage, perceived utility, and suggestions about CDSS tools, including the chief complaint section, question-and-answer-based workflow, and system-suggested diagnoses. Approximately 1,000 individuals participated in the survey, and the majority provided positive feedback, along with suggested corrections.

## Discussion

eSanjeevani 2.0 and subsequent versions are an example of AI-based CDSS solution working in the field of National health sector - the National Telemedicine service of MoHFW, Government of India, which has evolved into the world’s largest documented telemedicine implementation in the primary healthcare, facilitating 0.4-0.45 million consultations in a day. Thus, in busy Indian OPD having high patient volume, eSanjeevani is leveraging health & wellness center by effectively facilitating teleconsultation, through symptom entry algorithms helping to reach a diagnosis, recommendation, clinical decision support and overall improve patient care (dynamic present and future).

While CDSS can be classified into different types, there are two subtypes based on the AI methodologies used: knowledge-based CDSS (KB-CDSS) and data-based CDSS (DB-CDSS) [10]. Knowledge-based CDSSs use logical rules to produce outcomes in the form of recommendations to guide clinicians. There is always a source of knowledge in a knowledge-based CDSS, and instructions are drawn from literature, patient-centred protocols, guidelines, or expert knowledge. Data-based CDSS relies on machine learning or statistical pattern recognition techniques to simulate expert knowledge.

Diagnostic Decision Support Systems (DDSS), a subset of CDSS, have demonstrated varying degrees of success globally over the past decade. These systems aim to assist clinicians in formulating differential diagnoses based on patient symptoms and clinical data. In the United States, the DDSS platform Isabel has been successfully integrated into several hospital systems. By leveraging structured symptom entry and real-time algorithmic processing, Isabel improved diagnostic accuracy and clinician satisfaction in pediatric and internal medicine settings. A study reported significant diagnostic improvements and reduced cognitive errors among residents using DDSS tools integrated with Electronic Health Records (EHRs) [11]. In Mexico, DXplain, developed by Massachusetts General Hospital, was used in a randomized controlled trial involving 87 family medicine residents. The system provided superior diagnostic accuracy (84% vs. 74%) compared to unaided physician judgment. This success was attributed to DXplain’s capacity to synthesize symptom patterns and present comprehensive differential diagnoses along with justifications [12]. In India, Kunhimangalam et al. developed a fuzzy logic-based DDSS for diagnosing peripheral neuropathy, achieving 93% diagnostic accuracy across motor, sensory, and mixed neuropathies. The system used 24 structured symptom and test input fields and proved especially useful in primary care centers with limited access to neurologists [13]. Additionally, in Brazil, the Mediktor app, piloted for teletriage, showed good utility in prioritizing cases based on urgency using symptom-based input, demonstrating how DDSS can enhance efficiency and accessibility in community-based telemedicine [14].

While many developed nations have successfully implemented DDSS platforms within their digital health ecosystems, developing countries have been slower in adoption. This delay stems from infrastructural limitations, low digital literacy, fragmented healthcare delivery, and the need for localized linguistic and epidemiological adaptations. Thus, despite the theoretical potential, real-world deployment of AI-enabled DDSS in primary care and telemedicine settings remains rare in these contexts. Here, focus is first on structured data entry, then only DDSS can be built on.

Despite promising examples, DDSS have faced significant implementation barriers and performance-related setbacks. One major limitation is negative physician perception. In multiple settings, including large health systems in Canada and Germany, physicians expressed mistrust toward DDSS recommendations, citing poor transparency and loss of clinical autonomy. Furthermore, systems that require manual data input—without seamless EHR integration—faced resistance due to increased documentation burden.

Another common failure mode has been poor accuracy due to incomplete or unreliable data inputs. This was seen in the Babylon AI Triage and Diagnostic System in the UK. While initially praised for its user-friendly interface and AI-driven DDSS capabilities, Babylon faced backlash when real-world testing revealed inconsistencies in diagnosis and inappropriate triage advice. The system was eventually suspended in some areas and ceased key operations in 2023 due to concerns over clinical safety and financial viability [15]. A similar fate met Sensyne Health in the UK, which aimed to apply AI algorithms to NHS-collected datasets. Despite strong academic partnerships, the company failed to deliver clinically useful DDSS tools, struggling with data standardization and financial sustainability [16]. In the United States, IBM’s Watson for Oncology, once seen as a flagship DDSS tool, was ultimately withdrawn from several cancer centers after disappointing results. Despite initial enthusiasm, Watson failed to match expert diagnostic recommendations and over-relied on limited datasets, revealing the critical role of high-quality training data and contextual clinical judgment [17]. Lastly, Olive AI, which attempted to automate a range of clinical and administrative functions, including diagnosis, faced closure due to unsustainable development costs and low adoption by healthcare systems [18].

In this context, our prospective implementation study within the eSanjeevani 2.0 and next versions represents a landmark initiative. It leverages the established strengths of DDSS - standardized symptom repository, guided data collection, and differential diagnosis generation—while addressing known barriers to adoption in developing countries. Our innovation is rooted in the development of an AI CDSS with a structured PAF for symptomatology entry using SNOMED-CT terminology. This system was refined through three iterative phases of development and expert validation involving independent clinical decision-makers and in 14 languages. It not only enables structured, multilingual data entry but also optimizes real-time diagnostic recommendations tailored to Indian epidemiology. The successful integration and implementation of validated AI-CDSS PAF in eSanjeevani 2.0 and next versions allows feasibility in high volume teleconsultation. Moving forward requires continuous validation and feedback refinement for offering insights that can enhance its diagnostic accuracy, functionality and acceptance among physicians/CHOs in resource-constrained and high demand public health settings.

In many developing countries, creating standalone Personal Health Records (PHRs) and EHRs can be prohibitively costly and time-consuming. A more sustainable alternative is the use of an integrated Clinical Decision Support System (iCDSS)—a shared digital platform that enables both patients and providers to collaborate during clinical consultations. By incorporating physician-assist and patient-assist forms into a unified interface, iCDSS facilitates collaborative diagnosis and treatment decisions, improving consultation quality and efficiency. This model represents a meaningful shift towards patient-focused care, where the CDSS helps overcome the common barrier of “lack of information,” enabling patients to actively participate in their health decisions [19]. To take it further, this iCDSS should be utilised not only in teleconsultation, but also in the entire e-hospital system, including NextGen. As a result, CDSS-supported iCDSS tools become ideal enablers of shared decision-making, particularly in resource-limited settings where cost-effective solutions are essential for scaling quality healthcare. The present PAF used in eSanjeevani 2.0 can easily be used as iCDSS since it incorporates exactly what patient says in Indian languages. Only deployment remains for them in the real world; perhaps eSanjeevani 3.0 will deploy iCDSS for both patients and physicians, including NextGen implementation.

## Conclusions

By embedding this AI-CDSS into eSanjeevani’s national teleconsultation framework, India becomes the first developing country to pilot a government-supported, large-scale AI-enabled CDSS platform within routine digital health services. This model addresses linguistic diversity, low-resource settings, and the urgent need for iCDSS support in primary care, all without compromising clinical autonomy or patient safety. Thus, our study offers a replicable, scalable, and contextually grounded pathway for other low- and middle-income countries (LMICs) seeking to leapfrog traditional barriers and adopt AI-CDSS in telemedicine. It sets a global example of how resource-constrained settings can lead to the ethical, inclusive, and practical deployment of clinical AI tools. More importantly, a shared decision between patient and provider through AI-iCDSS is the closest hope for developing countries like India to launch future versions of eSanjeevani.

## Data Availability

It will be made available to others as required upon request for the corresponding author.

## DATA SHARING

It will be made available to others as required upon request for the corresponding author.

## ACKNOWLEDGEMENTS

Thanks to Wadhwani AI team for giving technical support, CDAC Mohali team for giving eSanjeevani data to use in this AI model and to deploy the AI solutions in the same platform. Also thanks to the eHealth unit of MOHFW for approving this implementation study and continuous guidance to AI Center of Excellence, AIIMS Rishikesh.

